# Availability, use and impact of workplace mental health supports during the COVID-19 pandemic in a Canadian cohort of healthcare workers

**DOI:** 10.1101/2023.12.12.23299862

**Authors:** Shannon Ruzycki, Anil Adisesh, Igor Burstyn, Quentin Durand-Moreau, France Labreche, Tanis Zadunayski, Nicola Cherry

## Abstract

The COVID-19 pandemic focused attention on workplace mental health (MH) supports for healthcare workers (HCWs).

**Methods:** HCWs in a Canadian cohort reported availability and use of workplace MH supports in October 2020, April 2021 and 2022. At recruitment (April-October 2020) they reported pre-pandemic MH. They completed the Hospital Anxiety and Depression Scale (HADS) at each contact. Availability and use of supports were examined by pandemic phase, workplace, work role and, for use, gender, age, pre-pandemic and current MH. Impact was assessed as MH in 2021/2022 following use in 2020.

**Results:** Reports of availability, use and HADS scores were obtained from 4400 HCWs working with patients. Access to MH supports increased during the pandemic, with 94% reporting access to some workplace support by 2022. Half the HCWs had at least one clinically significant HADS score during the pandemic. The proportion with high anxiety scores decreased from 29% to 24% as the pandemic progressed: proportions with high depression scores remained close to 10%. Those with a history of pre-pandemic or current mental ill-health formed the majority of HCWs using MH supports. 25% of those with high HADS scores did not use supports, with depressed males least likely to report use. HCWs using an Employment Assistance Program at the 2^nd^ contact had lower HADS scores at next follow-up but this was not sustained.

**Conclusion:** HCWs reported increasing availability and use of MH supports as the pandemic progressed but one in four of those with anxiety and, particularly, depression did not seek support.

## Introduction

During the early months of the COVID-19 pandemic there was concern about the effects on the mental health of healthcare work (Greenberg et al. 2020; Shanafelt et al. 2020). Early reports suggested that those working with patients known or suspected to be infected with the SARS-CoV-2 virus were anxious and depressed (Salari et al. 2020; Spoorthy et al. 2020; Hennein and Lowe 2020; Li et al. 2021; Zhang et al. 2021). Given the uncertainties about the length and intensity of the pandemic and the demands that would be made on healthcare workers (HCWs), organizations employing HCWs had to consider the appropriateness, availability and effectiveness of the mental health (MH) supports in place to minimize MH impacts of the pandemic. There was rather little empirical evidence to indicate which approaches would be valuable (Pollock et al. 2020; Zaçe et al. 2021). Before the pandemic there had been studies that evaluated mental health interventions for HCWs at the individual (Tamminga et al. 2023) or organizational level (Gray et al. 2019), without providing good evidence for specific interventions. The start of a pandemic was not an environment for the cluster randomized trials that might have resulted in clear evidence of benefit of workplace mental health programs, but a number of short-term interventions were assessed in controlled trials (Amsalem et al. 2022; Coifman et al. 2021; Fiol-DeRoque et al. 2021). There have been thoughtful reviews of support programs put in place for HCWs during the pandemic (Buselli et al. 2021; Villarreal-Zegarra et al. 2022; David et al. 2022; Härkänen et al. 2023) but again without specific recommendations as to the type of intervention, while recognizing the need both for better support with both employee and employer engagement and for more research.

The study reported here describes the availability, use and perceived effectiveness of individual level interventions reported by members of a Canadian cohort of HCWs employed in many organizations over four Canadian provinces. The aims were to examine differences in the MH supports offered through work by work roles and workplaces, and to see how these changed as the pandemic progressed; to examine the degree to which personal characteristics of the HCW, particularly previous or concurrent mental ill-health, may have influenced either use or neglect of available supports, and, finally, to consider whether, in the absence of randomization, the longitudinal data collection in this cohort study could provide any useful indication of the effect of use of supports on HCWs mental health during the pandemic.

## Background

This study was carried out among healthcare workers in Canada, where healthcare is free at the point of service, with delivery of healthcare organized by the province or territory. Long term care of those unable to live unassisted may be provided by for-profit organizations. Physicians are very largely paid on a fee-for service basis by the provincial health authority and, even if working in a public institution such as a hospital, are not employees. Health care organizations must comply with provincial health and safety regulations and, during the COVID-19 pandemic, followed provincial public health directions on matters such as quarantine and vaccination. Professional organizations, for example provincial medical associations, provided MH supports to complement those provided by the employer.

## Methods

We recruited a cohort of HCWs from four Canadian provinces (Alberta, British Columbia, Ontario and Quebec) close to the start of the COVID-19 pandemic and followed them for 24 months. The respondents completed online questionnaires, in English or French, at recruitment in the spring/summer of 2020 (Phase 1), in the late fall of 2020 (Phase 2), the spring of 2021 (Phase 3) and the spring/summer of 2022 (Phase 4). As described in more detail elsewhere (Cherry et al. 2023), HCWs were approached through their professional organizations within each province. Physicians (MDs) were recruited from Alberta, British Columbia, Ontario and Quebec, registered nurses and registered psychiatric nurses (RNs), licensed practical nurses (LPNs) and health care aides (HCAs) just from Alberta and personal support workers (PSWs) just from Ontario. At the recruitment questionnaire participants were asked about any history of mental ill-health and whether they had received any treatment for anxiety or depression in the 12 months before the start of the pandemic in Canada, in March 2020. They were also asked their age and the gender with which they identified and to confirm their work role (MD, RN, LPN, PSW, HCA).

At recruitment and at each subsequent follow-up the HCWs completed a series of questions about their workplace from which we coded, for each contact, whether any of their work was in a hospital, in the community, in a residential institution (such as a care-home or prison) or in the patient’s own home, with the possibility of combinations of workplaces, together with details of their work with patients. At each contact they were also asked to complete the Hospital Anxiety and Depression Scale (HADS) (Bjelland et al. 2002), giving an indication of anxiety and depression in the previous week. HADS comprises 14 questions (7 for anxiety and 7 for depression) scored 0-3 with resulting scores from 0-21 on each dimension. On both the anxiety and depression scales a score of 11 or greater may be interpreted as a degree of anxiety or depression of clinical significance (Zigmond and Snaith 1983). Although higher than the statistically optimal cut-point discussed elsewhere (Bjelland et al. 2002) a cut-point of 11 or greater would be expected to have the greatest specificity (Brennan et al. 2010).

In the second, third and fourth contacts they were also asked ‘Since the start of the pandemic has your employer, professional organization, union or other group offered support to help you cope with the mental health challenges of the pandemic’. For each of six supports they were asked whether the support had been offered and, if yes, whether they took advantage of it. On the 4^th^ questionnaire in 2022 (but not earlier) those who reported using the support were asked to rate how helpful the support had been, indicating this on a visual analogue scale, from 0 (not at all useful) to 100 (very useful).

The six items in the checklist were

- one-on-one support (online or in person) from a specialist counsellor, psychologist, or similar;
- one-on-one support from a colleague or peer nominated to do this;
- an online support group where you could discuss and ask questions;
- an online self-learning tool with advice about how to manage stress;
- a helpline with a number you could call if you were distressed;
- an Employee Assistance Program (EAP)

In addition, the respondent was asked if there were ‘any other mental health supports you would have valued during this time’ (with an open text option) and if, since the start of the pandemic, they had ‘discussed any mental health issues with a health professional (such as your family doctor) you accessed through channels outside work’.

### Statistical methods

Analysis was restricted to those working with patients at the time of completing the questionnaire at phase 2, 3 or 4. Multilevel logistic regression examined the reported availability of each type of MH support by workplace, work role and phase of the pandemic. Reported use was examined with the same model including in addition, age, gender and history of MH in the 12 months before the pandemic. The proportion of users considered a case of mental ill-health currently (HADS≥11) or pre-pandemic, was calculated for each type of support for each phase of the pandemic and adjusted odds of ‘caseness’ (current or pre-pandemic) estimated. Characteristics of those with high HADS scores who reported no use of any MH support were examined overall and stratified by gender. The lagged effect of use of a MH support at phase 2 on anxiety and depression at phase 3 and 4 was examined by multilevel logistic regression adjusting for pre-pandemic mental ill-health and phase 1 and 2 anxiety and depression scores.

To improve estimates of availability and use of supports ‘cumulative’ variables were constructed to indicate total reporting of availability and use up to each phase. Reports at phase 2 were taken as is, but the cumulative report at phase 3 also included reports at phase 2 and those at phase 4 reports on any of the phase 2-4 questionnaires. A similar process was used for discussion with health professionals outside work. All participants were assumed to have no-cost access to a health care professional outside work. As the cumulative estimates were not statistically independent, modelling that included indication of phase used reported rather than estimated access and use. Analysis was carried out in Stata 18 (StataCorp. 2023. Stata Statistical Software: Release 18. College Station, TX: StataCorp LLC). All p values quoted are two-sided.

### Ethical approval

Approval for each element of the study was given by the University of Alberta Heath Ethics Board (Pr000099700). The study was also reviewed and approved by Unity Health Toronto Research Ethics Board (REB# 20-298) for those elements coordinated locally for the Ontario participants. The research was carried out in accordance with the Declaration of Helsinki. All participants gave online written informed consent.

## Results

Of the 4964 HCWs recruited for the study 4400 of those working with patients reported the availability of workplace MH supports on at least one questionnaire, for a total of 11262 reports. The characteristics of these HCWs are given in Table 1. The majority were female, with nurses as the largest professional group. More than 20% reported having been treated for anxiety or depression in the 12 months before the start of the pandemic. The proportion whose HADS questionnaire indicated an anxiety case decreased from 29% to 24% as the pandemic progressed. The proportion whose score on the depression scale indicated a case was 10% at recruitment and remained at that level. Overall, 51% (2239/4400) of these HCWs working with patients had a HADS score suggesting a case of anxiety or depression on one or more of the four occasions the scale was completed during the pandemic.

**Table 1.** Characteristics of health care workers working with patients at the time of completing their last questionnaire (N = 4400).

|  |  | N | % |
| --- | --- | --- | --- |
| Gender: | Not female | 723 | 17.6 |
|  | Female | 3627 | 82.4 |
| Workplace: | Hospital | 2823 | 64.2 |
|  | Community | 1384 | 31.5 |
|  | Residential | 574 | 13.0 |
|  | Home care | 452 | 10.3 |
| Work role: | MD | 1355 | 30.8 |
|  | RN | 2721 | 61.8 |
|  | LPN | 66 | 1.5 |
|  | PSW | 184 | 4.2 |
|  | HCA | 74 | 1.7 |
| Completed: | Phase 2 | 4064 | 100.0 |
|  | Phase 3 | 3708 | 91.2 |
|  | Phase 4 | 3490 | 85.9 |
| Mental ill-health before pandemic | No | 3032 | 68.9 |
|  | Yes | 940 | 21.4 |
|  | Unknown | 428 | 9.7 |
| HADS anxiety *'case' | Phase 1 | 1119 | 29.1 |
|  | Phase 2 | 1100 | 27.1 |
|  | Phase 3 | 950 | 25.6 |
|  | Phase 4 | 840 | 24.1 |
| HADS depression *'case' | Phase 1 | 386 | 10.1 |
|  | Phase 2 | 400 | 9.8 |
|  | Phase 3 | 377 | 10.2 |
|  | Phase 4 | 348 | 10.0 |

\* % of those working with patients who completed the Hospital Anxiety and Depression Scale (HADS) with a score >10.
N=3814 (phase 1) 4064(phase 2) 3708 (phase 3)3490(phase 4)

Table 2 shows the reports of availability and use of supports as reported on the questionnaire together with ‘cumulative’ reports. At phase 4, 60% reported the availability of a specialist counsellor, for example, and 12% reported they had used such a resource. When earlier reports are included with those from phase 4, the proportions increase to 77% availability and 18% use. On either metric, specialist counsellors, a helpline if distressed and an employment assistance program were the supports most frequently reported to be available and a nominated peer support the least. The support most frequently used was an online stress tool, with use of a helpline if distressed having the fewest reports of use.

**Table 2.** Reported and cumulative availability and use of workplace mental health supports. Since the start of the pandemic has your employer offered supports (availability)? Did you take advantage of this (use)? Those working with patients at the time of completing the questionnaire.

|  | Phase | Availability |  |  |  | Use |  |  |  | N<br>questionnaires |
| --- | --- | --- | --- | --- | --- | --- | --- | --- | --- | --- |
|  |  | Reported |  | Cumulative |  | Reported |  | Cumulative |  |  |
|  |  | n | % | n | % | n | % | n | % |  |
| Specialist<br>counsellor | 2 | 1799 | 44.3 | 1799 | 44.3 | 252 | 6.2 | 252 | 6.2 | 4064 |
|  | 3 | 1961 | 52.9 | 2387 | 64.4 | 325 | 8.8 | 432 | 11.7 | 3708 |
|  | 4 | 2091 | 59.9 | 2703 | 77.4 | 428 | 12.3 | 635 | 18.2 | 3490 |
|  | Overall | 5851 | 52.0 | 6889 | 61.2 | 1005 | 8.9 | 1319 | 11.7 | 11262 |
| Nominated<br>peer<br>support | 2 | 610 | 15.0 | 610 | 15.0 | 203 | 5.0 | 203 | 5.0 | 4054 |
|  | 3 | 606 | 16.3 | 919 | 24.8 | 204 | 5.5 | 334 | 9.0 | 3708 |
|  | 4 | 532 | 15.2 | 1087 | 31.1 | 203 | 5.8 | 425 | 12.2 | 3490 |
|  | Overall | 1748 | 15.5 | 2616 | 23.2 | 610 | 5.4 | 962 | 8.5 | 11262 |
| Online<br>support<br>group | 2 | 1036 | 25.5 | 1036 | 25.5 | 295 | 7.3 | 295 | 7.3 | 4064 |
|  | 3 | 1077 | 29.0 | 1534 | 41.3 | 239 | 6.4 | 438 | 11.8 | 3708 |
|  | 4 | 750 | 21.5 | 1645 | 47.1 | 198 | 5.7 | 511 | 14.6 | 3490 |
|  | Overall | 2863 | 25.4 | 4215 | 37.4 | 732 | 6.5 | 1244 | 11.0 | 11262 |
| Online<br>stress tool | 2 | 1644 | 40.5 | 1644 | 40.5 | 428 | 10.5 | 428 | 10.5 | 4064 |
|  | 3 | 1736 | 46.8 | 2240 | 60.4 | 501 | 13.5 | 713 | 19.2 | 3708 |
|  | 4 | 1384 | 39.7 | 2353 | 67.4 | 462 | 13.2 | 860 | 24.6 | 3490 |
|  | Overall | 4764 | 42.3 | 6237 | 55.4 | 1391 | 12.4 | 2001 | 17.8 | 11262 |
| Helpline if<br>distressed | 2 | 2064 | 50.8 | 2064 | 50.8 | 81 | 2.0 | 81 | 2.0 | 4064 |
|  | 3 | 2125 | 57.3 | 2615 | 70.5 | 103 | 2.8 | 137 | 3.7 | 3708 |
|  | 4 | 1916 | 54.9 | 2784 | 80.1 | 117 | 3.4 | 204 | 5.8 | 3490 |
|  | Overall | 6105 | 54.2 | 7473 | 66.4 | 301 | 2.7 | 422 | 3.7 | 11262 |
| Employee<br>Assistance<br>Program | 2 | 2183 | 53.7 | 2183 | 53.7 | 185 | 4.6 | 185 | 4.6 | 4064 |
|  | 3 | 2094 | 56.5 | 2420 | 65.3 | 243 | 6.6 | 317 | 8.5 | 3708 |
|  | 4 | 1956 | 56.0 | 2510 | 71.9 | 346 | 9.9 | 477 | 13.7 | 3490 |
|  | Overall | 6233 | 55.3 | 7113 | 63.2 | 774 | 6.9 | 979 | 8.7 | 11262 |

The number of supports reported available and used by phase, workplace and job are shown in Table 3, again using cumulative reports. Overall, 10% of questionnaires reported no access to supports, with 63% reporting access to at least 3 of the 6 listed. The proportions reporting use of at least one support increased as the pandemic progress with, overall, 35% of questionnaires, as a cumulative estimate, including a report of some use. The cumulative estimated use of supports accessed outside work (33%) was very similar.

**Table 3.** Cumulative availability and use of work support and support outside work by phase, workplace and work role (N=11262 observations from 4400 healthcare workers working with patients).

|  |  | Number of workplace supports available |  |  |  |  |  | Use of any workplace supports |  | Any use of supports not accessed through work |  | N |
| --- | --- | --- | --- | --- | --- | --- | --- | --- | --- | --- | --- | --- |
|  |  | None |  | 1-2 |  | 3-6 |  |  |  |  |  |  |
|  |  | n | % | n | % | n | % | n | % | n | % |  |
| Phase | 2 | 690 | 17.0 | 1568 | 38.6 | 1806 | 44.4 | 955 | 23.5 | 946 | 23.3 | 4064 |
|  | 3 | 274 | 7.4 | 943 | 25.4 | 2491 | 67.2 | 1362 | 36.7 | 1263 | 34.1 | 3708 |
|  | 4 | 149 | 4.3 | 573 | 16.4 | 2768 | 79.3 | 1627 | 46.6 | 1528 | 43.8 | 3490 |
| Workplace* | Hospital | 705 | 9.7 | 2001 | 27.6 | 4535 | 62.6 | 2360 | 32.6 | 2351 | 32.5 | 7241 |
|  | Community | 411 | 9.9 | 1143 | 27.4 | 2616 | 62.7 | 1508 | 36.2 | 1351 | 32.4 | 4170 |
|  | Residential | 136 | 9.1 | 427 | 28.6 | 930 | 62.3 | 644 | 43.1 | 561 | 37.6 | 1493 |
|  | Homecare | 46 | 6.7 | 159 | 23.6 | 470 | 69.7 | 330 | 49.0 | 275 | 40.8 | 674 |
| Work role** | MD | 379 | 10.3 | 992 | 27.0 | 2306 | 62.7 | 1172 | 23.8 | 916 | 24.9 | 3677 |
|  | RN | 651 | 9.5 | 1857 | 27.2 | 4323 | 63.3 | 2437 | 28.0 | 2566 | 37.6 | 6831 |
|  | LPN | 12 | 7.2 | 52 | 31.1 | 103 | 61.7 | 73 | 43.7 | 70 | 41.9 | 167 |
|  | PSW | 53 | 12.9 | 119 | 29.0 | 239 | 58.2 | 184 | 44.8 | 123 | 29.9 | 411 |
|  | HCA | 18 | 10.2 | 64 | 36.4 | 94 | 53.4 | 78 | 44.3 | 62 | 35.2 | 176 |
| Overall | N | 1113 | 9.9 | 3084 | 27.4 | 7065 | 62.7 | 3944 | 35.0 | 3737 | 33.2 | 11262 |

\*Respondents could work in multiple workplaces
\*\*MD medical doctor; RN registered nurse; LPN licensed practical nurse; PSW personal support worker; HCA health care aide

Details of the supports reported as available are shown in Supplementary Materials (SM Table 1). Reported access to all types of support increased from phase 2 to phase 3 with that to specialist counsellors and EAPs increasing further at phase 4. There was rather little difference in access by place of work: those working in homecare reported less access to peer support but more to online stress tools. Those working in the community were most likely to report access to an online support group but least to an EAP. Differences were more marked by work role. Physicians were most likely to have access to nominated peer support and to an online support group but much less likely to report access to an EAP. HCAs were least likely to have access to a specialist counsellor or an EAP.

Reported use details are given in Supplementary Materials (SM Table 2) for those who reported that the workplace support was available to them. At all three phases support from a nominated peer was most used among those who had access. A helpline and the EAP were least used. Those who worked in a hospital were less likely to use each of the supports, with those working in residential or homecare the most likely. Physicians, followed by RNs, were least likely to use supports other than the helpline. The use of the support by gender and reported history of treatment or mental ill-health in the 12 months before the start of the pandemic are also shown in SM Table 2. Adjusted odds ratios, indicating likelihood of use, are given in Table SM3. Older workers reported less use of specialised counsellors, and EAPs but more of online stress tools. HCWs identifying as female were more likely to report use of each support, with the difference most marked for online tools. Those reporting pre-pandemic treatment for anxiety or depression were more likely to use each of the available workplace MH supports.

Those who used supports were asked, at phase 4, how useful they had found them (Table 4). Online stress tools and EAPs got the highest ratings, each with a median of 48 (on a scale from 0 – no use at all to 100 – very useful). Support from a nominated peer got the lowest ratings (a median of 18). There was rather little systematic difference in rating between the subgroups making up the cohort. Details are given in SM Table 4. Nurses gave higher ratings of usefulness to EAPs and a helpline than did physicians. Those working in residential institutions found the online support groups helpful and those working in a hospital, the online stress management tools. Older workers gave lower ratings to support from nominated peers and to online stress management tools.

**Table 4.**
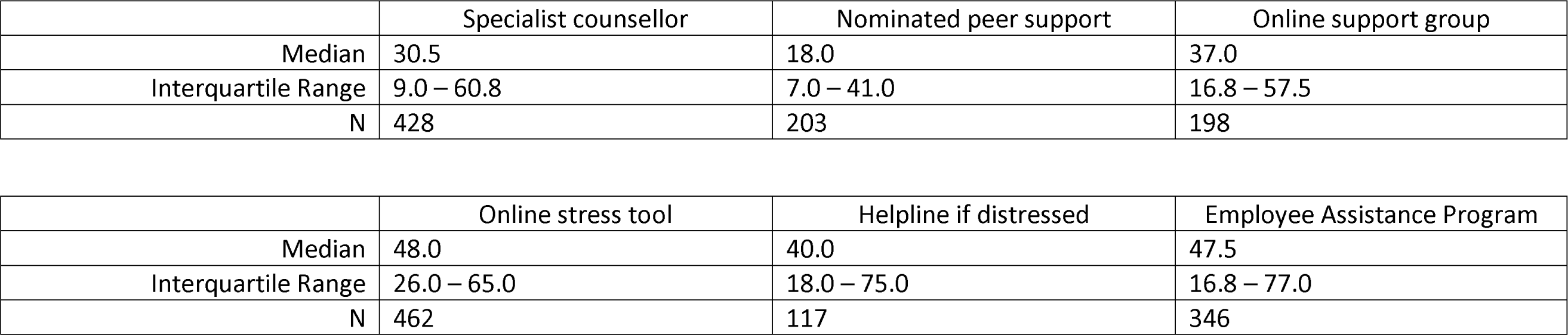
Rated usefulness of supports by those who reported use at Phase 4.

The proportions of those using each type of support who were a ‘case’ of anxiety or depression is shown in Table 5. Here respondents were considered a ‘case’ if they had reported treatment for anxiety or depression in the year before the pandemic or whose HADS scores (11 or greater) suggested, at the time of reporting use, clinically significant anxiety or depression. The majority of those using peer support, online support groups and online stress tools had no such pre-pandemic or current anxiety or depression, whereas a majority of those gaining access to a specialist counsellor (61%), or using an EAP (62%) or helpline (59%) were currently, or prior to the pandemic, unwell to a degree that would warrant assessment or intervention. As the pandemic progressed, the proportion of those seeking support who met the criterion for a ‘case’ decreased (with a decrease also in the associated adjusted OR) for all supports except use of an online support group, in which the proportion of cases stayed fairly constant and use of a helpline, where there was an increasing proportion of cases among those reporting use.

**Table 5.** Proportion of those using supports (where available) who were classified as a ‘case’ of anxiety or depression currently (from HADS) or pre-pandemic.

| Support used | Phase | N case | % | N use | OR* | 95% CI | P = |
| --- | --- | --- | --- | --- | --- | --- | --- |
| Specialist counsellor | 2 | 168 | 66.7 | 252 | 3.28 | 2.46 – 4.37 | <0.001 |
|  | 3 | 195 | 60.0 | 325 | 2.50 | 1.95 – 3.21 | <0.001 |
|  | 4 | 251 | 58.6 | 428 | 2.51 | 2.01 – 3.14 | <0.001 |
|  | Overall | 614 | 61.1 | 1005 | 5.51 | 3.74 – 8.12 | <0.001 |
| Nominated peer support | 2 | 89 | 43.8 | 203 | 1.27 | 0.86 – 1.86 | 0.225 |
|  | 3 | 97 | 47.5 | 204 | 1.85 | 1.25 – 2.71 | 0.002 |
|  | 4 | 80 | 39.4 | 203 | 1.35 | 0.89 – 2.06 | 0.160 |
|  | Overall | 266 | 43.6 | 610 | 2.25 | 1.22 – 4.15 | 0.009 |
| Online support group | 2 | 138 | 46.8 | 295 | 1.79 | 1.34 – 2.40 | <0.001 |
|  | 3 | 104 | 43.5 | 239 | 1.43 | 1.06 – 1.94 | 0.020 |
|  | 4 | 90 | 45.5 | 198 | 1.97 | 1.39 – 2.78 | <0.001 |
|  | Overall | 332 | 45.4 | 732 | 2.87 | 1.74 – 4.72 | <0.001 |
| Online stress tool | 2 | 218 | 50.9 | 428 | 1.83 | 1.45 – 2.32 | <0.001 |
|  | 3 | 254 | 50.7 | 501 | 1.55 | 1.24 – 1.93 | <0.001 |
|  | 4 | 214 | 46.3 | 462 | 1.53 | 1.21 – 1.94 | <0.001 |
|  | Overall | 686 | 49.3 | 1391 | 2.46 | 1.77 – 3.43 | <0.001 |
| Helpline if distressed | 2 | 46 | 56.8 | 81 | 1.91 | 1.20 – 3.05 | 0.006 |
|  | 3 | 59 | 57.3 | 103 | 1.95 | 1.30 – 2.95 | 0.001 |
|  | 4 | 73 | 62.4 | 117 | 2.77 | 1.86 – 4.12 | <0.001 |
|  | Overall | 178 | 59.1 | 301 | 3.75 | 1.96 – 7.16 | <0.001 |
| Employee Assistance Program | 2 | 125 | 67.6 | 185 | 2.52 | 1.82 – 3.50 | <0.001 |
|  | 3 | 152 | 62.6 | 243 | 2.02 | 1.52 – 2.67 | <0.001 |
|  | 4 | 200 | 57.8 | 346 | 1.96 | 1.54 – 2.50 | <0.001 |
|  | Overall | 477 | 61.6 | 774 | 2.58 | 1.78 – 3.72 | <0.001 |

\*Adjusted for gender, workplace, work role and age.

A summary was made for each participant by compiling all the data provided over each of the four phases. This (Table 6) allowed us to identify those whose HADS scores from phase 1 to phase 4 had ever suggested a level of anxiety or depression that would warrant assessment/intervention and their use of any supports at work or outside at any point in the pandemic (up to July 2022). There were only 271/4440 (6.2%) who, by the time of their last questionnaire, had not reported access to supports at work. Of note are 535/2114 (25%) who had high HADS scores and access to work supports but did not use either those or support from a health professional outside work. In a logistic regression analysis (Table 7) those with access to work supports were more likely to use support (at work or outside). HCWs identifying as female were more than twice as likely (OR=0.47) as non-female HCWs to seek help: 75% (1461/1940) of female HCWs who were ever a case based on HADS score used supports compared with 55% (165/299) of male or non-binary. Stratification by gender suggested that older female HCWs with anxiety or depression were less likely than younger ones to use supports, while among non-female HCWs older workers were more likely to report use. Among those not identifying as female, HCWs with depression (and not anxiety) were particularly unlikely to use supports: only 38% (11/29) of these reported using any support, at work or outside. Of the 18 not using supports (13 MD, 5 RNs, all identifying as male), five answered, on at least one questionnaire, the open-ended question about the additional help they would have valued. One MD, whose last questionnaire was at phase 2, reported that he would have liked access to all the supports listed but he was far to busy to use them, another MD commented at phase 4, that the health authority should have focused on the need to return to normalcy, two (one MD and one RN) commented on the lack of support from their – different – provincial government and two (one MD and one RN) suggested that paid time away would have been helpful.

**Table 6.**
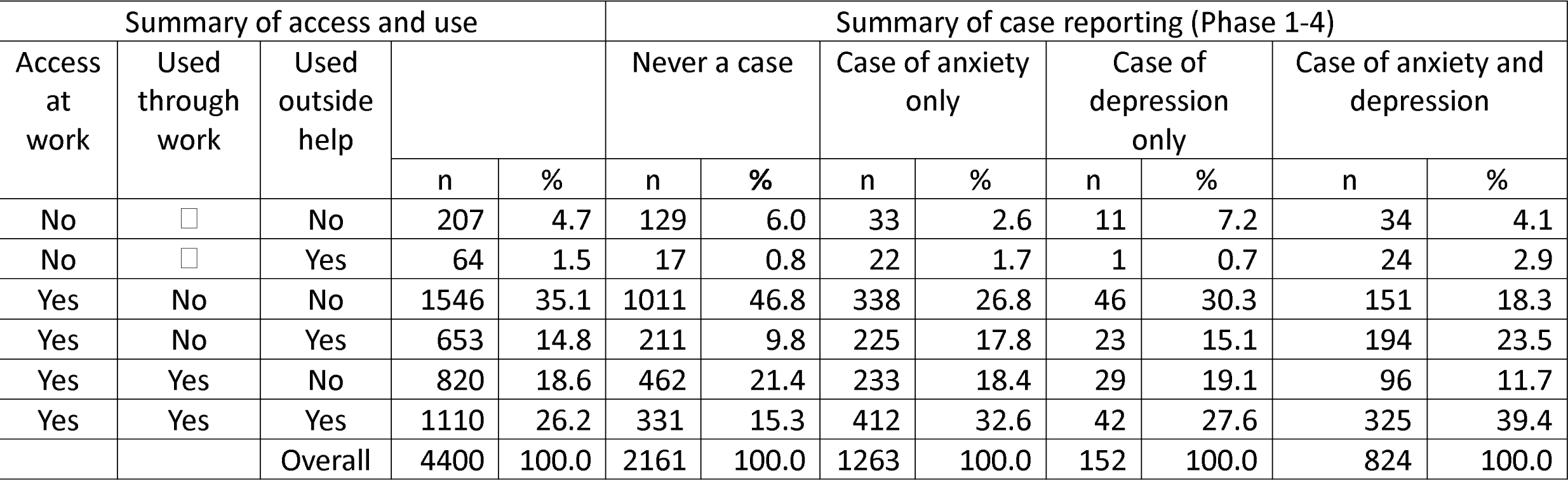
Summary of access, use and case status from HADS for phases 1-4. N=4400 healthcare workers.

| Summary of access and use |  |  |  |  | Summary of case reporting (Phase 1-4) |  |  |  |  |  |  |  |
| --- | --- | --- | --- | --- | --- | --- | --- | --- | --- | --- | --- | --- |
| Access at work | Used through work | Used outside help |  |  | Never a case |  | Case of anxiety only |  | Case of depression only |  | Case of anxiety and depression |  |
|  |  |  | n | % | n | % | n | % | n | % | n | % |
| No | ☐ | No | 207 | 4.7 | 129 | 6.0 | 33 | 2.6 | 11 | 7.2 | 34 | 4.1 |
| No | ☐ | Yes | 64 | 1.5 | 17 | 0.8 | 22 | 1.7 | 1 | 0.7 | 24 | 2.9 |
| Yes | No | No | 1546 | 35.1 | 1011 | 46.8 | 338 | 26.8 | 46 | 30.3 | 151 | 18.3 |
| Yes | No | Yes | 653 | 14.8 | 211 | 9.8 | 225 | 17.8 | 23 | 15.1 | 194 | 23.5 |
| Yes | Yes | No | 820 | 18.6 | 462 | 21.4 | 233 | 18.4 | 29 | 19.1 | 96 | 11.7 |
| Yes | Yes | Yes | 1110 | 26.2 | 331 | 15.3 | 412 | 32.6 | 42 | 27.6 | 325 | 39.4 |
|  |  | Overall | 4400 | 100.0 | 2161 | 100.0 | 1263 | 100.0 | 152 | 100.0 | 824 | 100.0 |

**Table 7.** Likelihood (odds ratio) of NOT using supports if ever a case of anxiety or depression.

|  |  | All (N = 2239) |  |  | Not female (N=299) |  |  | Female (N= 1940) |  |  |
| --- | --- | --- | --- | --- | --- | --- | --- | --- | --- | --- |
|  |  | OR | 95% CI | P= | OR | 95% CI | P= | OR | 95% CI | P= |
| Gender | Not female | 1.00 | — | — | — | — | — | — | — | — |
|  | Female | 0.47 | 0.36 to 0.63 | <0.001 | — | — | — | — | — | — |
| Age (continuous) x 10 years |  | 1.07 | 0.98 to 1.16 | 0.146 | 0.74 | 0.59 to 0.93 | 0.011 | 1.13 | 1.03 to 1.25 | 0.009 |
| Work role* | MD | 1.00 | — | — | 1.00 | — | — | 1.00 | — | — |
|  | RN | 0.81 | 0.64 to 1.03 | 0.091 | 0.58 | 0.32 to 1.06 | 0.075 | 0.82 | 0.63 to 1.07 | 0.137 |
|  | LPN | 0.70 | 0.32 to 1.52 | 0.369 | 1.00 | — | — | 0.78 | 0.35 to 1.70 | 0.530 |
|  | PSW | 1.16 | 0.72 to 1.86 | 0.535 | 3.70 | 0.66 to 20.86 | 0.138 | 1.04 | 0.62 to 1.72 | 0.895 |
|  | HCA | 0.79 | 0.36 to 1.75 | 0.565 | 1.00 | — | — | 0.84 | 0.38 to 1.88 | 0.676 |
| Access to work supports |  | 0.20 | 0.14 to 0.30 | <0.001 | 0.23 | 0.09 to 0.55 | 0.001 | 0.19 | 0.12 to 0.30 | <0.001 |
| Anxiety case |  | 1.00 | — | — | 1.00 | — | — | 1.00 | — | — |
| Depression case |  | 1.25 | 0.87 to 1.81 | 0.225 | 2.53 | 1.07 to 5.97 | 0.034 | 1.10 | 0.73 to 1.67 | 0.652 |
| Anxiety and depression case |  | 0.63 | 0.51 to 0.78 | <0.001 | 0.82 | 0.49 to 1.37 | 0.439 | 0.60 | 0.47 to 0.76 | <0.001 |

\*MD medical doctor; RN registered nurse; LPN licensed practical nurse; PSW personal support worker; HCA health care aide

Although it was evident that those who sought help had more anxiety and depression at the time of reporting use of supports, benefit from use of supports might be evident going forward. The final analysis (Table 8) examined the possibility of such a lagged effect. The question of interest here was whether anxiety or depression were less at phase 3 or 4 in those who had reported on phase 2 that they had taken advantage of any available workplace MH support since the beginning of the pandemic. This analysis took account not only of mental ill-health before the pandemic but also of HADS scores recorded in phase 1 and 2 together with other potential confounders (workplace, work role, gender, age and availability of the support). Those reporting use of more supports at phase 2 were somewhat less likely to be working with patients at phase 3 (but not at phase 4) and the analysis in Table 8 is adjusted for this. Overall, those who reported at phase 2 that they had used an EAP had somewhat lower anxiety and depression scores at phase 3 (median weeks phase 2 completion to phase 3 completion =29). For anxiety, but not depression, this was reflected in a lower likelihood of being a case (anxiety OR=0.57 95%CI 0.36 to 0.90 p=0.016; depression OR=0.93 95%CI 0.53 to 1.63 p=0.801). Table 8 also shows that use of supports at phase 2 was unrelated to anxiety or depression at phase 4 (median weeks phase 2 completion to phase 4 completion =81), with the reduction in anxiety and depression scores in those who reported EAP use seen at phase 3 no longer in evidence.

**Table 8.**
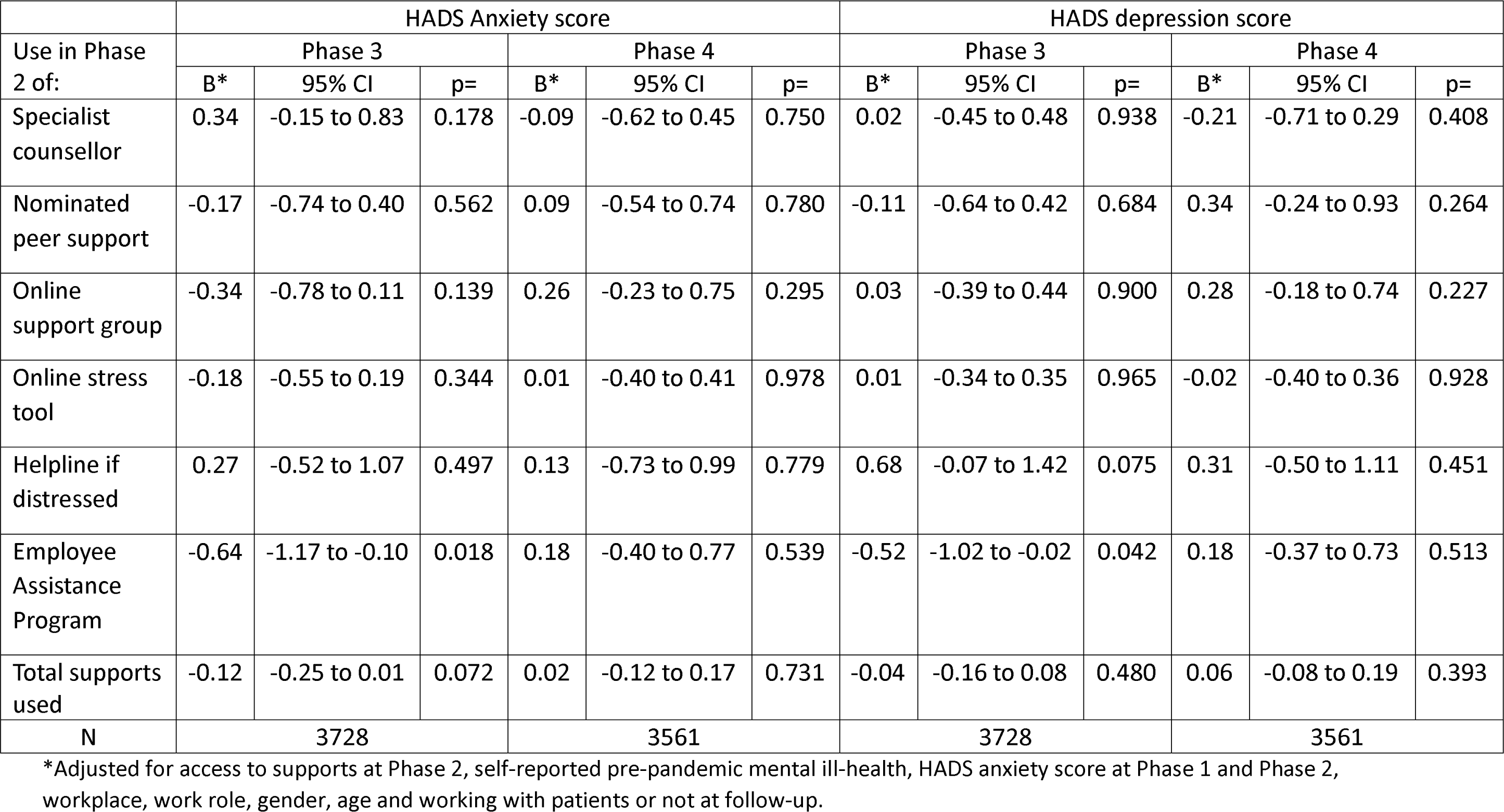
Impact on anxiety and depression at Phases 3 and 4 of use of mental health supports at Phase 2.

## Discussion

The great majority of HCWs reported access to some mental health support through their employment with an increase in reported access and use as the pandemic progressed. Overall, access to supports was similar across workplaces and work roles, with MDs reporting a different pattern of supports than other HCWs, with less access to EAPs and more to online support groups and nominated peer support. HCWs identifying as female were more likely to use available supports and older workers less likely. The median rating of usefulness was towards the negative end of the rating scale for each of the supports. Half the HCWs had a score suggesting clinically significant anxiety or depression at some point during the pandemic. The majority of those using supports had either a recent pre-pandemic history of treatment for mental ill-health or current indicators of anxiety or depression. One in four of those with scores suggesting a case of anxiety and/or depression during the pandemic did not report using any MH supports with more than 60% of men with isolated depression choosing not to do so. Those using an EAP had less anxiety and depression at a six-month (29 weeks) follow-up but this benefit was not in evidence in the longer term.

The strengths of this report include the large numbers of HCWs who joined the cohort and who, in repeated questionnaires, told us about their workplace and the supports it offered during the pandemic. The prospective study, with repeated contemporary assessment of anxiety and depression was also a strength, allowing assessment of change as the pandemic progressed. The data have a significant weakness in that we have no objective record of the supports actually offered but are dependent on the sometimes-inconsistent reports of the participant. We approximated a more complete response by construction of a cumulative record but this inconsistency in response would suggest an unknown degree of additional underreporting. Moreover, responses about the availability of supports may be biased by the respondent’s mental state: those deeply depressed, for example, may be reluctant to recognize that help is offered. As the pandemic progressed there may have been an increasing normalcy in discussion of MH supports and their use which may account for at least some of the increase in reported availability and use in later questionnaires. A further complexity arose from the multiple job locations at which HCWs might work. If an MD worked in a hospital emergency room, as a family doctor in the community and did sessions in an institution, we had no record of which worksite offered the supports they reported. We do not have good information on when use of a support actually occurred but can date it only by the questionnaire on which it was reported. It was not feasible in this study to validate through standardized clinical interviews the cut-point used to indicate clinically significant anxiety and depression as we have done elsewhere (Cherry et al. 2021) and from these data it is not evident that depression was necessarily related to the pandemic. Comparison with community referents does, however, show an excess risk of depression in these HCWs, rising as the pandemic progressed (Galarneau et al. 2023).

Even with confidence in data on the availability of supports, demonstration of their effectiveness would only be secure in a randomized intervention trial. As in any observational study there is a risk (here a strong probability) of ‘confounding by intention’. Those who seek help are, as seen here, those who are anxious or depressed and the decision to take some action may in itself be an indication of greater mental distress than is indicated by mental health scores alone. Insofar as this is the case, statistical adjustment will be incomplete.

A further limitation is that we know very little about the interventions that respondents reported. There are many online stress management tools, for example, and those within the study who reported using them may have had a wide range of experiences. Similarly, we do not know if the help from a specialist counsellor was face-to-face or online (probably the latter, to reduce infection risk), the experience of, or approach taken, by the counsellor or the frequency of sessions and length of support. Although it appears that use of an EAP was to a degree effective, we do not know what happened during that intervention. The finding of some benefit from an EAP intervention, at 6 months post reporting but not 18 months, is consistent with the conclusions of a recent Cochrane review on individual-level interventions, that stress management interventions may, in the short term, reduce stress in HCWs (Tamminga et al. 2023). It seems likely that organizational level changes, including management of workload, team building, improved communications and tangible workplace supports such as sick pay, while not well captured in these data, would help to reduce mental distress in future pandemics (Gray et al. 2019; David et al. 2022; Brand et al. 2017).

The demands of the pandemic on HCWs’ mental health were recognized early in 2020 together with the need to improve access to MH supports (Greenberg et al. 2020; Shanafelt et al. 2020), but there have been few reports of how these interventions evolved as the pandemic progressed, of their users’ perceptions or the gaps that became apparent. As such it is difficult to know how far the experiences of HCWs in this Canadian cohort relate to those working in very different health care systems. It seems certain that the many demands reported by these HCWs during the height of the pandemic were common worldwide. It would be helpful to know much more about the supports that were offered across different societies, as has been attempted (Villarreal-Zegarra et al. 2022; Muller et al. 2020; Byrne et al. 2023), but with only narrative assessment of the impacts of the interventions. Convincing arguments have been made of the need to provide organizational supports to prevent harm (through decreasing workforce demands and improving tangible supports such as childcare, for example), together with peer support and self-care training, rather than simply providing support to those whose mental health conditions have become manifest (David et al. 2022; Byrne et al. 2023). Nevertheless, during future times of crisis, those responsible for the mental health of HCWs will again be looking for clear indications of the strategies to adopt as well as the at-risk groups (older women, men with depressive conditions), who may fall through the cracks. The design and execution of evaluative studies (as a randomized trial or an interrupted time series) to evaluate MH supports in quieter times would provide the strongest evidence to inform practice when employers are reaching out for ways to reduce the toll on HCWs caught up in a developing societal threat.

## Funding

Seed funding was obtained from the College of Physicians and Surgeons of Alberta. Grant funding was obtained from the Canadian Institutes of Health Research (Funding Reference number 173209). This funding was extended by a grant from the Canadian Immunology Task Force

### Ethics approval and informed consent

Approval for each element of the study was given by the University of Alberta Heath Ethics Board (Pr000099700). The study was also reviewed and approved by Unity Health Toronto Research Ethics Board (REB# 20-298) for those elements coordinated locally for the Ontario participants. The research was carried out in accordance with the Declaration of Helsinki. All participants gave online written informed consent.

## Conflict of interest

All authors declare no conflicts of interest. Data availability statement

The data underlying this article will be shared on reasonable request to the corresponding author.

## Data Availability

The data underlying this article will be shared on reasonable request to the corresponding author.

