## Supplementary materials for "Availability, use and impact of workplace mental health supports during the COVID-19 pandemic in a Canadian cohort of healthcare workers"

SM Table 1. Reported availability of each type of mental health support by phase, workplace and work role in those with 1 on 1 patient care

SM Table 2. Use of each type of mental health support (in healthcare workers reporting access) by phase, workplace, work role, gender and self-reported mental ill-health in the year before the pandemic.

SM Table 3. Use of each type of mental health support (in those with access): multivariate multilevel logistic regression adjusting for gender, age and previous mental ill-health

SM Table 4. Rated usefulness of supports by those who reported use at Phase 4 with coefficients from a multilevel multivariable linear regression

SM Table 1. Reported availability of each type of mental health support by phase, workplace and work role in those with 1 on 1 patient care (N=11262 questionnaires from 4400 healthcare workers). Odds ratios (OR) from multilevel multivariable logistic regression

|  |  | **Specialist counsellor** | | | | | **Nominated peer support** | | | | | **Online support group** | | | | |
| --- | --- | --- | --- | --- | --- | --- | --- | --- | --- | --- | --- | --- | --- | --- | --- | --- |
|  |  | N | % | OR | 95% CI | P= | N | % | OR | **95% CI** | P= | N | % | OR | 95% CI | P= |
| Phase | 2 | 1805 | 44.3 | 1.0 | ̶ | ̶ | 611 | 15.0 | 1.0 | ̶ | ̶ | 1038 | 25.5 | 1.0 | ̶ | ̶ |
|  | 3 | 1963 | 52.8 | 1.6 | 1.5 to 1.8 | **<0.001** | 609 | 16.4 | 1.2 | 1.0 to 1.3 | **0.047** | 1078 | 29.0 | **1.3** | **1.1 to 1.4** | **<0.001** |
|  | 4 | 2091 | 59.9 | 2.6 | 2.3 to 2.9 | **<0.001** | 532 | 15.2 | 1.0 | 0.9 to 1.2 | 0.675 | 750 | 21.5 | **0.7** | **0.6 to 0.8** | **<0.001** |
| Workplace | Hospital | 3720 | 51.3 | 0.9 | 0.7 to 1.1 | 0.160 | 1140 | 15.7 | 0.9 | 0.8 to 1.2 | 0.605 | 1758 | 24.3 | 1.0 | 0.9 to 1.3 | 0.623 |
|  | Community | 2216 | 53.0 | 1.1 | 0.9 to 1.3 | 0.401 | 652 | 15.6 | 0.9 | 0.7 to 1.1 | 0.228 | 1312 | 31.4 | **1.5** | **1.2 to 1.7** | **<0.001** |
|  | Residential | 751 | 50.3 | 0.9 | 0.8 to 1.1 | 0.574 | 237 | 15.9 | 1.1 | 0.8 to 1.4 | 0.605 | 403 | 27.0 | **1.2** | **1.0 to 1.5** | **0.025** |
|  | Homecare | 361 | 53.6 | 1.0 | 0.7 to 1.3 | 0.898 | 86 | 12.8 | 0.6 | 0.4 to 0.9 | **0.017** | 161 | 23.9 | 1.3 | 0.9 to 1.7 | 0.108 |
| Work role | *MD | 1999 | 54.3 | 1.0 | ̶ | ̶ | 796 | 21.6 | 1.0 | ̶ | ̶ | 1437 | 39.0 | 1.0 | ̶ | ̶ |
|  | RN | 3503 | 51.2 | 0.9 | 0.7 to 1.0 | 0.080 | 834 | 12.2 | 0.4 | 0.3 to 0.5 | **<0.001** | 1261 | 18.4 | **0.3** | **0.2 to 0.3** | **<0.001** |
|  | LPN | 85 | 50.9 | 0.8 | 0.5 to 1.5 | 0.531 | 16 | 9.6 | 0.3 | 0.1 to 0.6 | **0.002** | 26 | 15.6 | **0.2** | **0.1 to 0.4** | **<0.001** |
|  | PSW | 205 | 49.9 | 0.8 | 0.5 to 1.2 | 0.269 | 75 | 18.2 | 1.0 | 0.6 to 1.7 | 0.999 | 113 | 27.5 | **0.5** | **0.3 to 0.7** | **<0.001** |
|  | HCA | 67 | 38.1 | 0.4 | 0.2 to 0.6 | **0.001** | 31 | 17.6 | 0.7 | 0.3 to 1.3 | 0.271 | 29 | 16.5 | **0.2** | **0.1 to 0.4** | **<0.001** |
|  |  | **Online stress tool** | | | | | **Helpline if distressed** | | | | | **Employee Assistance program** | | | | |
|  |  | N | % | OR | 95% CI | P= | N | % | OR | 95% CI | P= | N | % | OR | 95% CI | P= |
| Phase | 2 | 1649 | 40.5 | 1.0 | ̶ | ̶ | 2069 | 50.8 | 1.0 | ̶ | ̶ | 2186 | 53.7 | 1.0 | ̶ | ̶ |
|  | 3 | 1739 | 46.8 | **1.4** | **1.3 to 1.6** | **<0.001** | 2127 | 57.3 | **1.4** | **1.3 to 1.6** | **<0.001** | 2096 | 56.4 | **1.3** | **1.1 to 1.5** | **<0.001** |
|  | 4 | 1384 | 39.6 | 0.9 | 0.8 to 1.1 | 0.378 | 1916 | 54.9 | **1.2** | **1.1 to 1.4** | **<0.001** | 1957 | 56.0 | **1.4** | **1.2 to 1.6** | **<0.001** |
| Workplace | Hospital | 2986 | 41.2 | 1.0 | 0.9 to 1.2 | 0.760 | 3918 | 54.1 | 1.1 | 1.0 to 1.4 | 0.100 | 4113 | 56.8 | **1.4** | **1.1 to 1.8** | **0.003** |
|  | Community | 1841 | 44.0 | **1.3** | **1.1 to 1.5** | **<0.001** | 2367 | 56.6 | **1.3** | **1.1 to 1.5** | **<0.001** | 2020 | 48.3 | 1.2 | 1.0 to 1.5 | 0.064 |
|  | Residential | 659 | 44.1 | 1.1 | 0.9 to 1.3 | 0.222 | 799 | 53.5 | 1.1 | 1.0 to 1.4 | 0.125 | 830 | 55.6 | 1.2 | 0.9 to 1.5 | 0.198 |
|  | Homecare | 322 | 47.8 | **1.6** | **1.2 to 2.0** | **0.001** | 372 | 55.2 | **1.4** | **1.1 to 1.9** | **0.007** | 407 | 60.4 | 0.9 | 0.6 to 1.2 | 0.435 |
| Work role | *MD | 1354 | 36.8 | 1.0 | ̶ | ̶ | 2099 | 57.0 | 1.0 | ̶ | ̶ | 963 | 26.1 | 1.0 | ̶ | ̶ |
|  | RN | 3090 | 45.2 | **1.7** | **1.5 to 2.0** | **<0.001** | 3659 | 53.5 | **0.9** | **0.8 to 1.0** | **0.043** | 4860 | 71.1 | **34.8** | **26.8 to 45.1** | **<0.001** |
|  | LPN | 74 | 44.3 | 1.6 | 0.9 to 2.8 | 0.078 | 77 | 46.1 | **0.6** | **0.3 to 1.0** | **0.033** | 104 | 62.3 | **16.1** | **7.1 to 36.3** | **<0.001** |
|  | PSW | 177 | 43.1 | 1.1 | 0.8 to 1.7 | 0.500 | 193 | 47.0 | **0.5** | **0.4 to 0.8** | **0.001** | 220 | 53.5 | **10.7** | **6.1 to 18.9** | **<0.001** |
|  | HCA | 77 | 43.8 | 1.4 | 0.8 to 2.4 | 0.213 | 84 | 47.7 | 0.6 | 0.4 to 1.0 | 0.063 | 92 | 52.3 | **10.0** | **4.8 to 20.8** | **<0.001** |

*MD medical doctor; RN registered nurse; LPN licensed practical nurse; PSW personal support worker; HCA health care aide
SM Table 2. Use of each type of mental health support (in healthcare workers reporting access) by phase, workplace, work role, gender and self-reported mental ill-health in the year before the pandemic.

| Factor | Specialist counsellor | | | Nominated peer support | | | Online support group | | | Online stress tool | | | Helpline if distressed | | | Employee Assistance Program | | |
| --- | --- | --- | --- | --- | --- | --- | --- | --- | --- | --- | --- | --- | --- | --- | --- | --- | --- | --- |
|  | n | N | % | n | N | % | n | N | % | n | N | % | n | N | % | n | N | % |
| Phase |  |  |  |  |  |  |  |  |  |  |  |  |  |  |  |  |  |  |
| 2 | 252 | 1805 | 14.0 | 203 | 611 | 33.2 | 296 | 1038 | 28.5 | 430 | 1649 | 26.1 | 81 | 2069 | 3.9 | 185 | 2186 | 8.5 |
| 3 | 326 | 1963 | 16.6 | 204 | 609 | 33.5 | 239 | 1078 | 22.2 | 502 | 1739 | 28.9 | 103 | 2127 | 4.8 | 243 | 2096 | 11.6 |
| 4 | 428 | 2091 | 20.5 | 203 | 532 | 38.2 | 198 | 750 | 26.4 | 462 | 1384 | 33.4 | 117 | 1916 | 6.1 | 346 | 1957 | 17.7 |
| Workplace | |  |  |  |  |  |  |  |  |  |  |  |  |  |  |  |  |  |
| Hospital | 591 | 3720 | 15.9 | 324 | 1140 | 28.4 | 401 | 1758 | 22.8 | 787 | 2986 | 26.4 | 193 | 3918 | 4.9 | 489 | 4113 | 11.9 |
| Community | 370 | 2216 | 16.7 | 227 | 652 | 34.8 | 355 | 1312 | 27.1 | 548 | 1841 | 29.8 | 119 | 2367 | 5.0 | 249 | 2020 | 12.3 |
| Residential | 164 | 751 | 21.8 | 127 | 237 | 53.6 | 149 | 403 | 37.0 | 251 | 659 | 38.1 | 54 | 799 | 6.8 | 107 | 830 | 12.9 |
| Homecare | 88 | 361 | 24.4 | 56 | 86 | 65.1 | 62 | 161 | 38.5 | 126 | 322 | 39.1 | 29 | 372 | 7.8 | 77 | 407 | 18.9 |
| Work role | |  |  |  |  |  |  |  |  |  |  |  |  |  |  |  |  |  |
| *MD | 273 | 1999 | 13.7 | 135 | 796 | 17.0 | 343 | 1437 | 23.9 | 316 | 1354 | 23.3 | 115 | 2099 | 5.5 | 100 | 963 | 10.4 |
| RN | 633 | 3503 | 18.1 | 399 | 834 | 47.8 | 326 | 1261 | 25.9 | 915 | 3090 | 29.6 | 149 | 3659 | 4.1 | 607 | 4860 | 12.5 |
| LPN | 22 | 85 | 25.9 | 6 | 16 | 37.5 | 7 | 26 | 26.9 | 35 | 74 | 47.3 | 3 | 77 | 3.9 | 19 | 104 | 18.3 |
| PSW | 58 | 205 | 28.3 | 52 | 75 | 69.3 | 47 | 113 | 41.6 | 84 | 177 | 47.5 | 24 | 193 | 12.4 | 43 | 220 | 19.5 |
| HCA | 20 | 67 | 29.9 | 18 | 31 | 58.1 | 10 | 29 | 34.5 | 44 | 77 | 57.1 | 10 | 84 | 11.9 | 5 | 92 | 5.4 |
| Gender |  |  |  |  |  |  |  |  |  |  |  |  |  |  |  |  |  |  |
| Not female | 116 | 1045 | 11.1 | 70 | 378 | 18.5 | 120 | 620 | 19.4 | 127 | 681 | 18.6 | 46 | 1088 | 4.2 | 68 | 681 | 10.0 |
| Female | 890 | 4814 | 18.5 | 540 | 1374 | 39.3 | 613 | 2246 | 27.3 | 1267 | 2091 | 31.0 | 255 | 5024 | 5.1 | 706 | 5558 | 12.7 |
| Mental ill-health in year before pandemic | | | | | |  |  |  |  |  |  |  |  |  |  |  |  |  |
| No | 543 | 4139 | 13.1 | 421 | 1319 | 31.9 | 481 | 2076 | 23.2 | 859 | 3268 | 26.3 | 166 | 4335 | 3.8 | 409 | 4115 | 9.9 |
| Yes | 367 | 1281 | 28.6 | 128 | 305 | 42.0 | 188 | 593 | 31.7 | 406 | 1098 | 37.0 | 106 | 1290 | 8.2 | 280 | 1535 | 18.2 |
| Unknown | 96 | 439 | 21.9 | 61 | 128 | 47.7 | 64 | 197 | 32.5 | 129 | 406 | 31.8 | 29 | 487 | 6.0 | 85 | 589 | 14.4 |
| N |  | 5859 |  |  | 1752 |  |  | 2866 |  |  | 4772 |  |  | 6112 |  |  | 6239 |  |

*MD medical doctor; RN registered nurse; LPN licensed practical nurse; PSW personal support worker; HCA health care aide

SM Table 3. Use of each type of mental health support (in those with access): multivariate multilevel logistic regression adjusting for gender, age and previous mental ill-health

| Factor | Specialist counsellor | | | Nominated peer support | | | Online support group | | | Online stress tool | | | Helpline if distressed | | | Employee Assistance Program | | |
| --- | --- | --- | --- | --- | --- | --- | --- | --- | --- | --- | --- | --- | --- | --- | --- | --- | --- | --- |
|  | OR | 95% CI | P | OR | 95% CI | P | OR | 95% CI | P | OR | 95% CI | P | OR | 95% CI | P | OR | 95% CI | P |
| Phase |  |  |  |  |  |  |  |  |  |  |  |  |  |  |  |  |  |  |
| 2 | 1.0 | ̶ | ̶ | 1.0 | ̶ | ̶ | 1.0 | ̶ | ̶ | 1.0 | ̶ | ̶ | 1.0 | ̶ | ̶ | 1.0 | ̶ | ̶ |
| 3 | **1.4** | **1.1 - 1.9** | **0.011** | 1.1 | 0.7 - 1.6 | 0.729 | **0.7** | **0.5 - 0.9** | **0.019** | **1.2** | **1.0 - 1.5** | **0.041** | **1.7** | **1.1 - 2.7** | **0.012** | **1.9** | **1.4 - 2.5** | **<0.001** |
| 4 | **2.6** | **2.0 - 3.5** | **<0.001** | **1.7** | **1.1 - 2.6** | **0.017** | 0.9 | 0.7 - 1.3 | 0.666 | **1.7** | **1.3 - 2.1** | **<0.001** | **2.5** | **1.6 - 3.8** | **<0.001** | **4.5** | **3.3 - 6.1** | **<0.001** |
| Workplace | |  |  |  |  |  |  |  |  |  |  |  |  |  |  |  |  |  |
| **Hospital** | **0.6** | **0.4 - 0.9** | **0.016** | **0.4** | **0.2 - 0.8** | **0.008** | 0.9 | 0.6 - 1.4 | 0.642 | 0.9 | 0.6 - 1.2 | 0.388 | 1.5 | 0.8 - 2.8 | 0.156 | 0.7 | 0.5 -1.2 | 0.193 |
| Community | 0.9 | 0.6 - 1.3 | 0.463 | 1.0 | 0.6 - 1.7 | 0.922 | 1.2 | 0.8 - 1.8 | 0.305 | 1.1 | 0.8 - 1.4 | 0.626 | 1.3 | 0.8 - 2.3 | 0.298 | 1.1 | 0.8 - 1.6 | 0.590 |
| Residential | 1.1 | 0.7 - 1.7 | 0.693 | 1.5 | 0.8 - 2.8 | 0.248 | **2.0** | **1.2 - 3.1** | **0.005** | **1.4** | **1.0 - 2.0** | **0.034** | 1.5 | 0.8 - 2.8 | 0.168 | 1.1 | 0.7 - 1.7 | 0.788 |
| Homecare | 1.0 | 0.6 - 1.9 | 0.892 | 1.5 | 0.5 - 4.4 | 0.491 | 1.4 | 0.6 - 2.9 | 0.419 | 0.8 | 0.5 - 1.3 | 0..330 | 1.2 | 0.5 - 2.8 | 0.677 | 1.4 | 0.8 - 2.5 | 0.231 |
| Work role | |  |  |  |  |  |  |  |  |  |  |  |  |  |  |  |  |  |
| *MD | 1.0 | ̶ | ̶ | 1.0 | ̶ | ̶ | 1.0 | ̶ | ̶ | 1.0 | ̶ | ̶ | 1.0 | ̶ | ̶ | 1.0 | ̶ | ̶ |
| RN | 1.2 | 0.8 - 1.7 | 0.477 | **10.7** | **6.0 -19.2** | **<0.001** | 1.0 | 0.7 -1.5 | 0.947 | 1.4 | 1.0 - 1.9 | 0.032 | **0.5** | **0.3 - 1.0** | **0.042** | 1.0 | 0.6 -1.6 | 0.925 |
| LPN | 1.3 | 0.4 - 4.0 | 0.619 | 2.9 | 0.4 - 20.9 | 0.287 | 0.7 | 0.1 - 4.1 | 0.679 | **3.7** | **1.5 - 9.4** | **0.006** | 0.7 | 0.1 - 5.1 | 0.684 | 1.9 | 0.5 - 7.0 | 0.324 |
| PSW | 2.2 | 0.9 - 5.3 | 0.090 | **24.7** | **6.4 - 94.9** | **<0.001** | 1.9 | 0.7 - 5.5 | 0.235 | **4.3** | **2.1 - 8.9** | **<0.001** | **6.6** | **1.8 - 24.4** | **0.004** | 2.0 | 0.7 - 5.6 | 0.215 |
| HCA | **4.5** | **1.2 - 16.5** | **0.025** | **14.7** | **3.1 - 70.2** | **0.001** | 1.7 | 0.4 -7.7 | 0.490 | **6.5** | **2.6 - 16.6** | **<0.001** | **6.6** | **1.3 - 34.4** | **0.025** | 0.3 | 0.1 - 1.4 | 0.120 |
| Gender |  |  |  |  |  |  |  |  |  |  |  |  |  |  |  |  |  |  |
| Not female | 1.0 | ̶ | ̶ | 1.0 | ̶ | ̶ | 1.0 | ̶ | ̶ | 1.0 | ̶ | ̶ | 1.0 | ̶ | ̶ | 1.0 | ̶ | ̶ |
| Female | **1.9** | **1.1 - 3.1** | **0.015** | 1.7 | 0.9 - 3.3 | 0.094 | **1.9** | **1.2 - 3.0** | **0.011** | **2.3** | **1.5 - 3.4** | **<0.001** | 1.7 | 0.8 - 3.5 | 0.142 | 1.4 | 0.8 - 2.6 | 0.227 |
| Mental ill-health in the 12months before the pandemic | | | | | |  |  |  |  |  |  |  |  |  |  |  |  |  |
| No | 1.0 | ̶ | ̶ | 1.0 | ̶ | ̶ | 1.0 | ̶ | ̶ | 1.0 | ̶ | ̶ | 1.0 | ̶ | ̶ | 1.0 | ̶ | ̶ |
| Yes | **5.6** | **3.8 - 8.2** | **<0.001** | **1.9** | **1.1 - 3.4** | **0.024** | **2.0** | **1.3 - 3.0** | **0.001** | **2.0** | **1.5 - 2.7** | **<0.001** | **3.8** | **2.3 - 6.5** | **<0.001** | **3.2** | **2.2 - 4.8** | **<0.001** |
| Unknown | 2.2 | 1.3 - 3.8 | 0.005 | 1.5 | 0.7 - 3.1 | 0.332 | 1.9 | 1.0 - 3.7 | 0.043 | 1.3 | 0.8 - 1.9 | 0.293 | 2.7 | 1.3 - 5.7 | 0.007 | 1.6 | 0.9 -2.7 | 0.116 |
| Age (continuous) | |  |  |  |  |  |  |  |  |  |  |  |  |  |  |  |  |  |
| Years/10 | **0.7** | **0.6 - 0.8** | **<0.001** | 0.9 | 0.8 - 1.1 | 0.408 | 1.0 | 0.9 - 1.2 | 0.667 | **1.3** | **1.1 - 1.4** | **<0.001** | 0.9 | 0.7 - 1.1 | 0.169 | **0.7** | **0.6 - 0.8** | **<0.001** |

*MD medical doctor; RN registered nurse; LPN licensed practical nurse; PSW personal support worker; HCA health care aide

SM Table 4. Rated usefulness of supports by those who reported use at Phase 4 with coefficients from a multilevel multivariable linear regression

|  |  | **Specialist counsellor** | | | **Nominated peer support** | | | **Online support group** | | |
| --- | --- | --- | --- | --- | --- | --- | --- | --- | --- | --- |
|  |  | β | 95 % CI | P= | β | 95 % CI | P= | β | 95 % CI | P= |
| Gender: female |  | -2.57 | -12.70 – 7.57 | 0.619 | -9.53 | -21.24 – 2.18 | 0.110 | 1.70 | -9.14 – 12.55 | 0.757 |
| Age (continuous) |  | -1.27 | -4.09 – 1.55 | 0.376 | **-3.32** | **-6.18 – -0.46** | **0.023** | 1.48 | -1.80 – 4.75 | 0.375 |
| Workplace | Hospital | -0.47 | -9.53 – 8.60 | 0.919 | -0.09 | -9.09 – 8.91 | 0.985 | 6.99 | -2.49 – 16.47 | 0.147 |
|  | Community | -3.56 | -12.87 – 5.75 | 0.452 | 2.17 | -6.28 – 10.62 | 0.613 | 1.64 | -8.34 – 11.63 | 0.746 |
|  | Residential | 1.00 | -8.41 – 10.42 | 0.834 | 2.44 | -7.07 – 11.95 | 0.614 | **15.21** | **4.63 – 25.78** | **0.005** |
|  | Homecare | -2.37 | -12.49 – 7.75 | 0.645 | -3.79 | -14.50 – 6.93 | 0.487 | 8.78 | -5.79 – 23.34 | 0.236 |
| Work role* | MD | 0.00 | – | – | 0.00 | – | – | 0.00 | – | – |
|  | RN | 8.26 | -0.18 – 16.70 | 0.055 | 0.74 | -8.28 – 9.76 | 0.871 | -0.36 | -10.07 – 9.35 | 0.942 |
|  | LPN | 4.17 | -17.30 – 25.64 | 0.703 | 7.29 | -17.32 – 31.90 | 0.560 | -5.40 | -45.62 – 34.83 | 0.792 |
|  | PSW | 7.71 | -10.03 – 25.45 | 0.394 | 3.39 | -13.34 – 20.12 | 0.690 | **-23.79** | **-44.62 – -2.96** | **0.025** |
|  | HCA | -2.13 | -26.24 – 21.98 | 0.862 | 12.36 | -6.60 – 31.32 | 0.200 | 0.69 | -28.14 – 29.52 | 0.962 |
| Pre-pandemic mental ill-health | No | 0.00 | – | – | 0.00 | – | – | 0.00 | – | – |
|  | Yes | 0.12 | -6.57 – 6.82 | 0.971 | 0.54 | -7.72 – 8.80 | 0.898 | 1.20 | -7.92 – 10.31 | 0.796 |
|  | Unknown | 3.73 | -6.14 – 13.61 | 0.458 | -1.57 | -14.29 – 11.15 | 0.808 | 10.39 | -3.89 – 24.68 | 0.153 |
|  |  | **Online stress tool** | | | **Helpline if distressed** | | | **Employee Assistance Program** | | |
|  |  | β | 95 % CI | P= | β | 95 % CI | P= | β | 95 % CI | P= |
| Gender: female |  | -1.60 | -10.61 – 7.42 | 0.728 | -14.07 | -32.54 – 4.40 | 0.134 | -6.93 | -20.29 – 6.42 | 0.308 |
| Age (continuous) |  | **-3.37** | **-5.42 – -1.31** | **0.001** | 1.39 | -4.25 – 7.03 | 0.626 | -1.52 | -4.79 – 1.74 | 0.359 |
| Workplace | Hospital | **6.89** | **0.05 – 13.73** | **0.048** | 9.31 | -7.23 – 25.84 | 0.267 | 4.64 | -6.17 – 15.45 | 0.399 |
|  | Community | 3.09 | -3.54 – 9.73 | 0.360 | 5.21 | -11.41 – 21.83 | 0.535 | -5.51 | -16.43 – 5.41 | 0.321 |
|  | Residential | -3.08 | -10.14 – 3.98 | 0.391 | 10.14 | -6.43 – 26.71 | 0.228 | -0.07 | -11.44 – 11.30 | 0.990 |
|  | Homecare | 1.41 | -5.87 – 8.68 | 0.704 | 11.36 | -8.14 – 30.86 | 0.251 | 4.14 | -6.74 – 15.02 | 0.455 |

| Work role* | MD | 0.00 | – | – | 0.00 | – | – | 0.00 | – | – |
| --- | --- | --- | --- | --- | --- | --- | --- | --- | --- | --- |
|  | RN | -1.35 | -7.99 – 5.28 | 0.689 | **23.39** | **7.95 – 38.83** | **0.003** | **16.22** | **4.99 – 27.45** | **0.005** |
|  | LPN | -9.78 | -25.89 – 6.32 | 0.233 | -22.71 | -91.07 – 45.64 | 0.511 | 24.83 | -0.88 – 50.55 | 0.058 |
|  | PSW | -8.59 | -21.80 – 4.61 | 0.202 | 23.30 | -9.42 – 56.02 | 0.161 | -0.89 | -20.83 – 19.04 | 0.930 |
|  | HCA | -11.38 | -26.28 – 3.52 | 0.134 | -5.47 | -70.99 – 60.05 | 0.869 | -9.17 | -55.96 – 37.61 | 0.700 |
| Pre-pandemic mental ill health | No | 0.00 | – | – | 0.00 | – | – | 0.00 | – | – |
|  | Yes | 3.59 | -1.93 – 9.10 | 0.202 | -6.66 | -20.35 – 7.03 | 0.337 | 3.53 | -4.12 – 11.18 | 0.364 |
|  | Unknown | 10.00 | 1.70 – 18.30 | 0.018 | 10.05 | -12.71 – 32.81 | 0.383 | 3.95 | -7.18 – 15.09 | 0.485 |

*MD medical doctor; RN registered nurse; LPN licensed practical nurse; PSW personal support worker; HCA health care aide
